# A novel functional assay based on patient-derived endometrial organoids and blastoids predicts the success of embryo transfer

**DOI:** 10.64898/2026.01.09.26343806

**Authors:** Elissa Tjahjono, Madeline Kaye, George M Grunert, Madhuri Pingle, Jack Crain, Subodh Chauhan, Ye Zhu, M. Cecilia Guerra, Scott Novich, Yael Katz, Aryeh Warmflash

## Abstract

Successful embryo implantation requires complex interactions between the embryo and the endometrium. Improvements in embryo testing have led to increased success rates for embryo transfer following in vitro fertilization, however, even with euploid embryos under ideal conditions, failure occurs in more than 30% of cases. Methods for diagnosing and improving endometrial function are currently lacking. Here we developed a functional test (“Simbryo FX”) for the ability of the endometrium to support embryo implantation. We collected endometrial biopsy samples from over 100 patients, and used these to grow endometrial organoids. We interacted these organoids with blastoids, pluripotent stem cell based models of the human blastocyst, and measured the hCG produced by the blastoids as well as the degree to which the blastoids invaded the organoids. We found that both of these measures correlated with clinical outcomes, and that combining them allowed us to predict the likelihood of failure in the next embryo transfer with high specificity. Thus, interacting patient-derived endometrial organoids with blastoids represents a promising approach to evaluating endometrial function among patients preparing for embryo transfer.

## Introduction

Despite improvements in clinical practice and embryo testing, success rates for embryo transfer following in vitro fertilization remain around 60%, and many patients undergo repeated embryo transfers without success^1^. While some have suggested that three or more unsuccessful transfers with euploid embryos are diagnostic of a condition known as recurrent implantation failure (RIF)^2^, others have suggested that most failures are due to stochastic factors and are not indicative of underlying pathology^3,4^. Current clinical recommendations are for up to six embryo transfers with euploid embryos^5^.

Several tests are available that measure aspects of the endometrium or uterine environment to diagnose conditions which may contribute to RIF including endometriosis, endometritis, and microbiome dysregulation^6–8^. However, while these conditions may affect fertility, they are not incompatible with pregnancy, and these tests do not provide insight into endometrial function. Receptivity assays performed in a mock cycle measure whether the endometrium is in the receptive phase so that adjustments can be made to embryo transfer timing^9,10^. Recent studies have questioned whether receptivity timing provides benefit for patients without a previous history of failure^11^. There is no available test to directly assess endometrial function.

Recent advances in in vitro culture of three dimensional tissue models, known as organoids, as well as pluripotent stem cell (PSC)-based models of embryos, open the possibility of directly modeling the interaction between endometrium and embryo. Endometrial organoids maintain the native structure and composition of the uterine epithelium and can be made to mimic the menstrual cycle through treatment with estrogen and progesterone^12–14^. Blastoids are models of the blastocyst created from naive pluripotent stem cells that contain all three lineages of the blastocyst: the epiblast, primitive endoderm, and trophectoderm^15–17^, and are capable of interacting specifically with endometrial cells when they are in a receptive state^15,18^. The specific interaction between blastoids and endometrial cells models early events in uterine implantation including the penetration of cells from the blastoid through the layer of endometrial epithelium and the production of hCG by the blastoid.

In this study, we developed a functional assay for endometrial function based on the interaction between endometrial organoids and blastoids (“Simbryo FX”). We collected over 100 endometrial biopsy samples from patients undergoing evaluation for infertility and analyzed gene expression in the primary samples and patient-derived organoids, and performed an assay in which we interacted the organoids with blastoids. While transcriptomic measurements did not correlate with clinical outcomes, metrics of the interaction between blastoids and organoids showed correlation with both retrospective data on the number of previous failed embryo transfers and prospective data on the outcome of the next transfer cycle for each patient. Taken together, our data show that measuring the interaction between blastoids and patient-derived organoids is a functional test of endometrial function that can be used to predict the likelihood of success in embryo transfer.

## Results

### Primary sample gene expression does not correlate with clinical outcomes

We aimed to study whether gene expression measurements of the endometrium, those from endometrial biopsy-derived organoids, or a functional assay testing blastoid adhesion to endometrial organoids could be predictive of clinical outcomes. As we were interested in whether these assays could be diagnostic of issues with the endometrium, we intentionally recruited a sample which overrepresented patients who had previous transfer failures as compared to the general population of patients undergoing embryo transfer (Figure S1A). In order to participate, patients were required to have at least one euploid embryo with intention to transfer the embryo within the study timeframe. All patients who participated were scheduled to undergo an endometrial biopsy as part of their routine care prior to enrolling in the study. Consistent with the idea that a fraction of these patients have endometrial issues, we found the likelihood of success in the upcoming transfer of a euploid embryo decreased strongly with the number of previous failures (Figure S1B). A portion of each biopsy was transferred to our laboratory and grown into organoids following standard procedures. Upon receipt, we split each sample into two parts, with one part used to grow organoids and the other part used to extract RNA in order to measure the transcriptome using RNA sequencing.

We compared outcomes in the clinic with expression of a panel of genes previously proposed to form a signature of receptivity^19^ and found that none showed a correlation with clinical outcomes (Figure 1A, S2A). More globally, we restricted the analysis to the 2000 most highly variable genes and performed PCA. While reliable data on the timing of the biopsy relative to the menstrual cycle was not available for all patients, a subset of patients were undergoing a biopsy for a window of implantation test, which involved a mock cycle with the biopsy taken at defined time, five days after beginning progesterone exposure. We found that the first principle component strongly correlated with whether patients were undergoing this test (Figure 1C), indicating that the menstrual cycle was the dominant influence on gene expression. None of the other components had a readily interpretable meaning and none correlated with outcomes (Figure 1B, S1D). Finally, we examined whether any genes were differentially expressed between patients who were successful in their next embryo transfer, defined as progressing to clinical pregnancy, and those who were not, defined as showing evidence of neither biochemical nor clinical pregnancy. We found that all genes showed less than two-fold expression difference between these categories (Figure 1D). We also examined whether any of the genes used in the endometrial receptivity array (ERA)^9^ to predict the window of implantation correlated with clinical outcomes and found that they did not (Figure S2B). Taken together, these data suggest that transcriptomics of the primary sample cannot be used to predict clinical outcomes.

**Figure 1.**
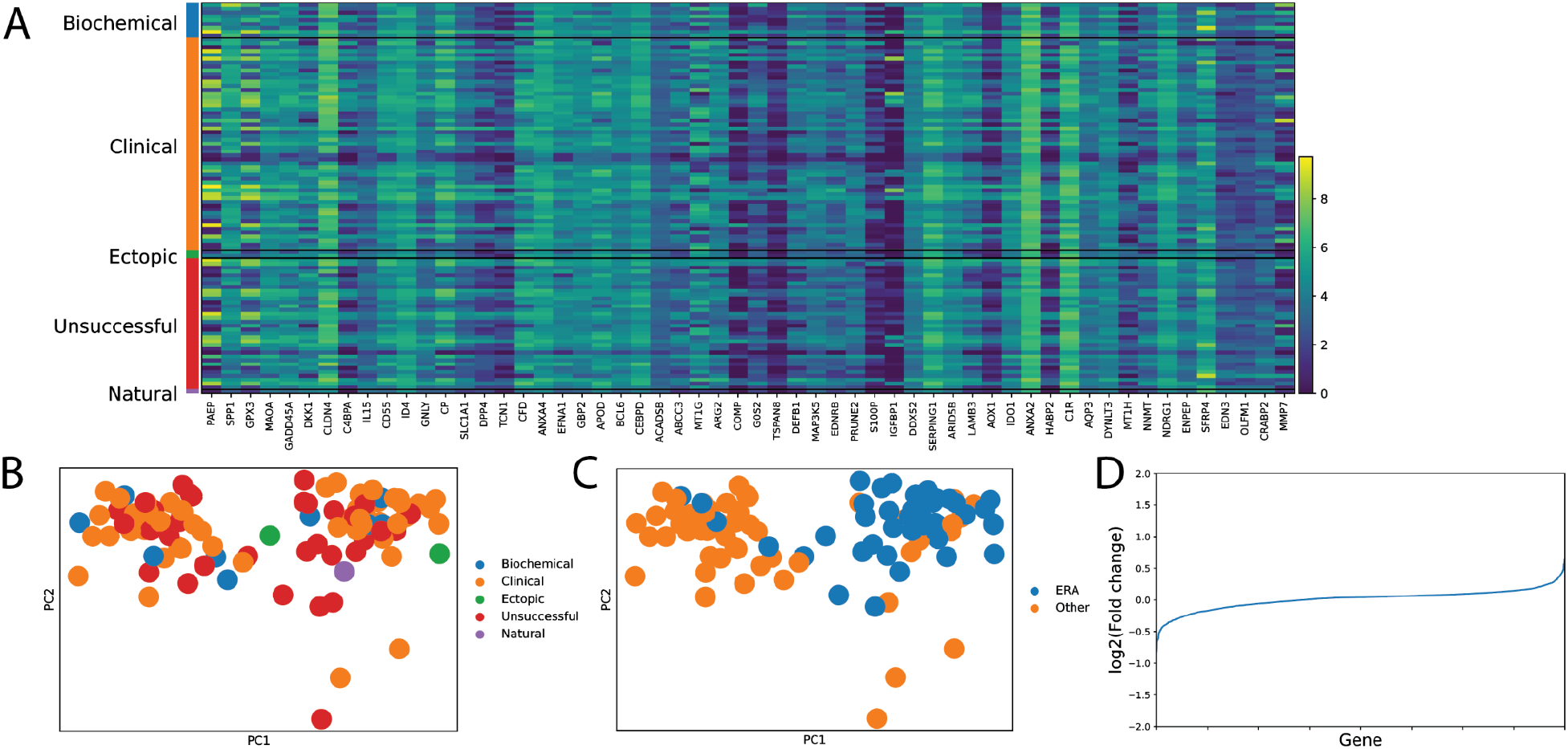
Primary sample transcriptomics are not predictive of clinical outcomes. (A) Heatmap showing expression of genes proposed to be a signature of endometrial receptivity with clinical outcomes indicated on the left. (B) PCA was run on the 2000 most highly variable genes and PC1 and PC2 were plotted with points color coded by clinical outcomes. (C) As in (B) but points were color coded by whether the biopsy was taken for a window of implantation test (ERA). (D)Fold changes (log2(unsuccessful/successful) of the 2000 most highly variable genes. All fold changes have magnitude less than 2.

### Patient-derived organoids are hormone responsive but organoid gene expression does not predict clinical outcomes

We created organoids from patient biopsies using standard procedures^12,13^. Organoids could be maintained in the lab for at least 10 passages and expressed markers of endometrium including E-CAD, MUC1, and SOX17 (Figure S3A). We treated organoids with a cycle of hormones which is typically used to prepare the endometrium for embryo transfer and includes two days of estrogen followed by five days of estrogen and progesterone. Upon hormone treatment, we observed a characteristic shift in morphology, with a reduction in cell density as organoid growth ceased (Figure S3B, top) and a thickening of the organoid layer (Figure S3B, bottom), consistent with what has been previously reported^13^.

We carried out this hormonal stimulation on all patient-derived organoids in parallel with mock-treated controls and performed RNA sequencing to gauge the response of each patient’s organoids to the hormones that prepare the endometrium for embryo transfer. All patient-derived organoids responded to hormone treatment, although the response varied by one to two orders of magnitude between the strongest and weakest responders depending on the gene (Figure 2A,B, Figure S4A). We examined whether these responses correlated with clinical outcomes, but did not find any relationship between response strength and the clinical outcome for any of the responsive genes we studied. More globally, performing PCA on the matrix of gene fold changes for each patient and gene also did not show any patterns that correlated with clinical outcome (Figure S4B). Importantly, the signature of cycle timing which was seen in the gene expression data from the primary biopsies (Figure 1C) was no longer present in the organoids (Figure S4B) indicating that the timing of biopsy collection did not influence the organoids. Thus, we found that organoids from all patients were hormone responsive but that the degree of response was not predictive of clinical outcomes.

**Figure 2.**
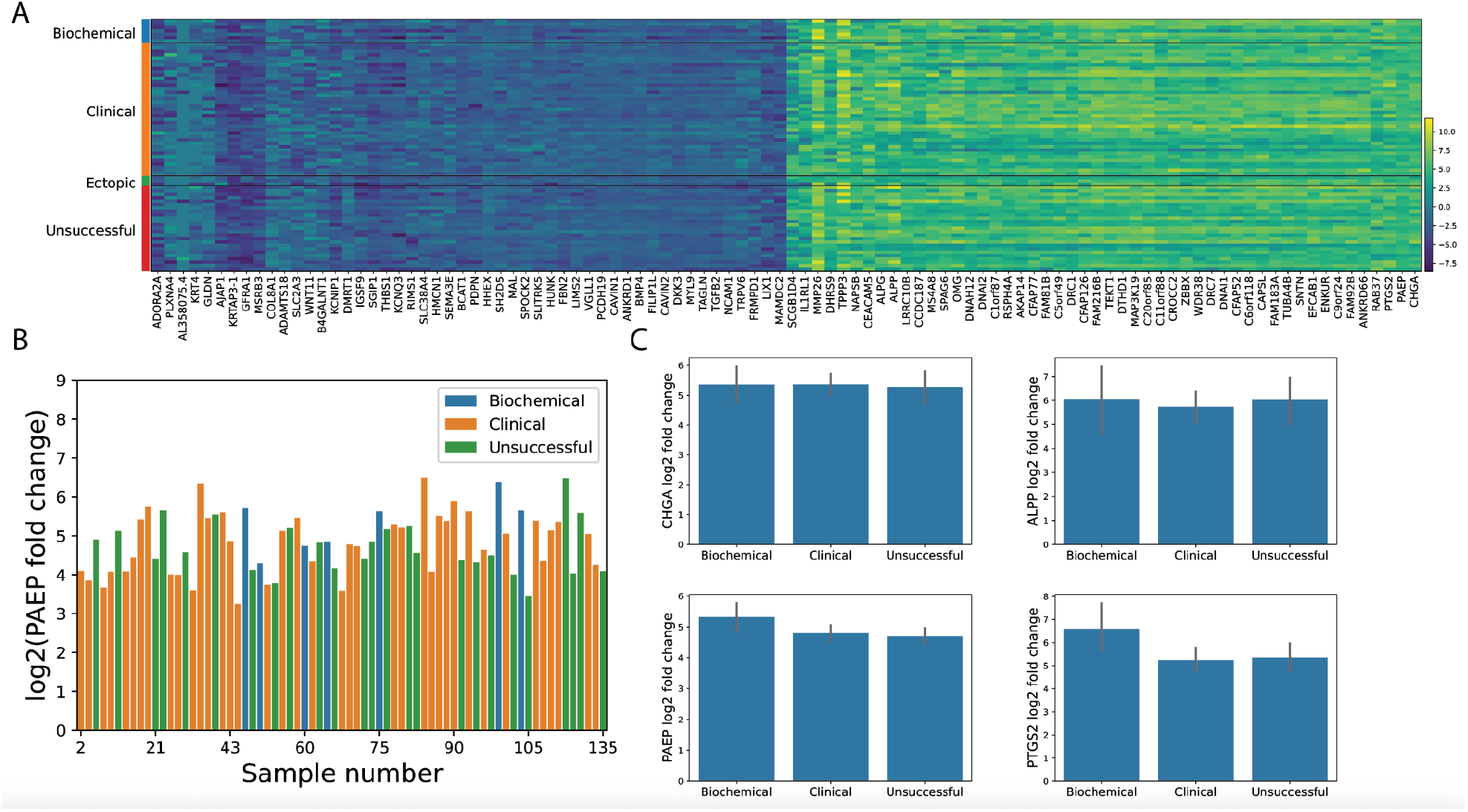
Patient-derived organoids are hormone responsive and the degree of response does not correlate with clinical outcomes. (A) Heatmap of the 50 most upregulated and 50 most downregulated genes between hormone-treated and control samples. Samples are grouped by clinical outcome which is indicated on the left. The color scale on the heatmap shows log2 of the fold change for each gene and sample. (B) PAEP expression fold change for each sample. Sample bars are color coded by clinical outcomes. (C) Average fold change as a function of clinical outcome for 4 progesterone responsive genes.

### Generation of naive pluripotent stem cells and blastoids for use in an implantation model

We next sought to create an assay to directly measure whether a patient’s endometrium could support the initial stages of embryo implantation. We used blastoids, stem-cell based models of the early human embryo, which have been shown to be capable of interacting with endometrial cells in vitro^15^. Blastoids are created from naive state pluripotent stem cells. We reset the primed state human pluripotent stem cell line ESI017 to this state using established methods^20,21^, and grew the resulting naive hPSCs in PXGL media. We analyzed gene expression by qPCR and found that cells reset to the naive state showed strong upregulation of naive markers including DNMT3L, DPPA3, DPPA5, and KLF17, while retaining expression of the general pluripotency markers OCT4, SOX2, and NANOG, and repressing expression of the primed-state specific markers SFRP2 and ZIC2 (Figure S5A). We performed differentiation to either trophectoderm (TE)^21^ or primitive endoderm (PE)^22^, and found upregulation of specific markers in the each case: CGB, GATA2, GATA3, KRT7, and SDC1 for TE, and GATA4, GATA6, NID2, and PDGRFa for PE (Figure S5B,C). Thus, the reset cells could be differentiated to represent all three lineages of the preimplantation embryo.

We generated blastoids according to an established protocol^15,18^ but optimized several parameters including the BSA batch and concentration, plate coating protocol, the concentrations and treatment times of the small molecules PD035921, A83-01, and LPA, and the number of passages in the naive state prior to blastoid generation (see methods for details). In particular, we found that even after cells had been reset and adopted naive morphology and markers, they needed to be grown in naive conditions for approximately 30 passages before routinely producing a high yield of blastoids (Figure S6A). Following this adaptation period, cells routinely produced a high yield of high quality blastoids with embryo-like morphology and containing all three lineages (Figure S6B).

### An optimized functional assay based on hCG production from blastoids

We next developed an optimized assay based on combining blastoids and endometrial organoids. In initial testing, we found that blastoids readily stick to glass or plastic tissue culture dishes and subsequently produce high levels of hCG, mimicking some aspects of the initial stages of implantation even without the presence of endometrial cells (Figure S7). Based on previous literature, we reasoned that although blastoids may interact non-specifically with tissue culture materials, they would not interact with unprimed endometrial cells. We also found that if endometrial cells were overgrown, they would block hCG production whether or not they were prepared with hormones. This raised the challenge that with too few endometrial cells, any exposed portions of the culture surface could lead to non-specific production of hCG, while with too many, hCG production would be blocked altogether.

In order to circumvent this issue and to standardize the patient-derived organoids as much as possible, we grew the organoid cells on micropatterns to create open layers of defined size and shape before introducing the blastoids. We found an optimal protocol was to grow the cells as organoids, perform hormone stimulation of the organoids as above until the final two days, and then dissociate the organoids to single cells which were then plated on micropatterns. Following culture on micropatterns for the final two days of hormone stimulation, blastoids were introduced and allowed to interact with the micropatterned organoids for an additional two days. At the conclusion of this time period, the media was collected for measurement of hCG levels by ELISA and the cultures were fixed for imaging. We found that endometrial cells on micropatterns showed stronger progesterone-dependent responses as measured by immunofluorescence for the progesterone target gene PAEP (Figure 3A) and induced robust production of hCG from blastoids even at high densities where production of hCG in standard culture was minimal (Figure 3B). Thus, this optimized protocol allowed us to measure the degree to which each patient’s endometrial cells were capable of supporting blastoid attachment and hCG production.

**Figure 3.**
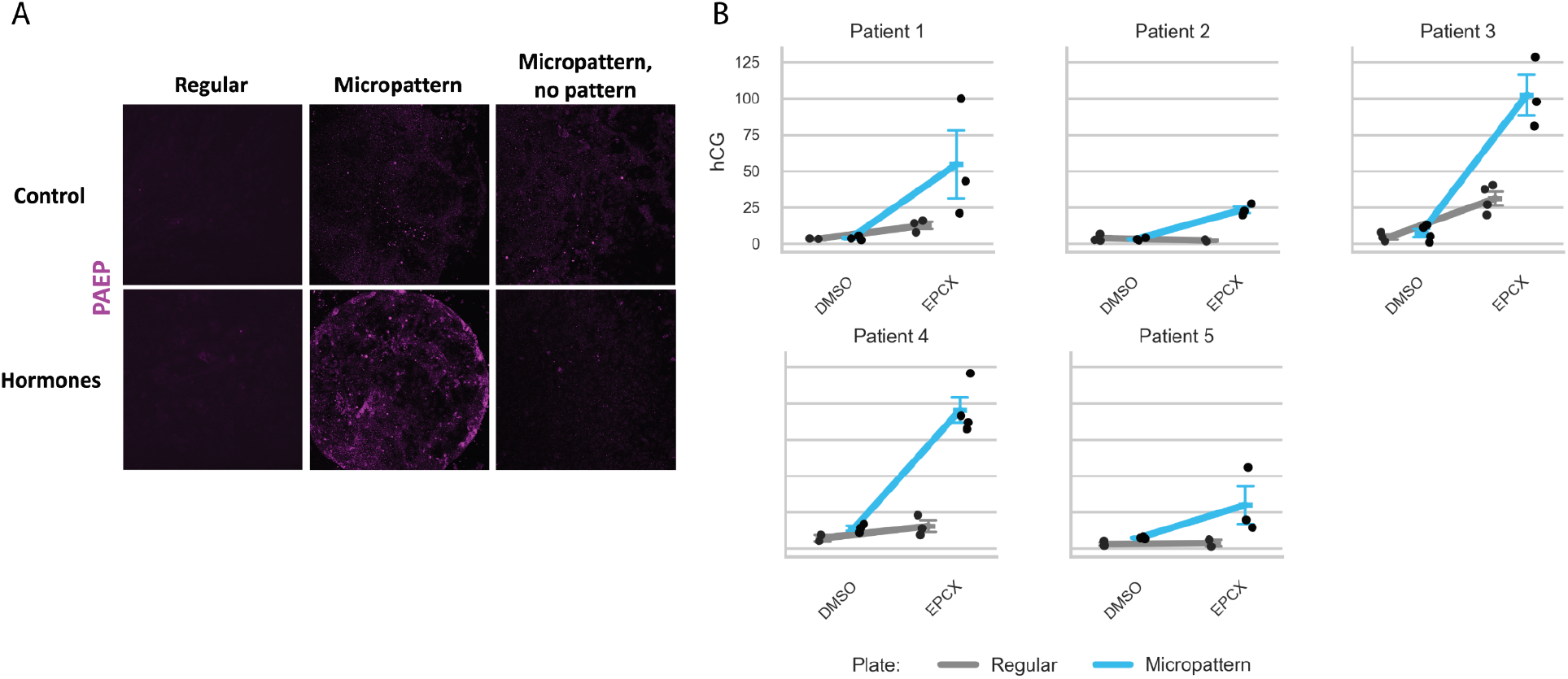
An optimized assay with blastoids and organoids for studying implantation. (A) Images of immunostaining for the progesterone responsive protein PAEP in regular or micropatterned culture. “Micropattern, no pattern” indicates the use of a micropatterned plate but in an experiment where the cells adhered non-specifically outside the patterns, showing that the strong PAEP response is specific to patterned colonies. (B) Comparison in hCG production from blastoids combined with endometrial organoid cells either on micropatterns or in standard culture for five different patients.

### hCG production from the blastoid-organoid assay correlates with clinical outcomes

We ran this assay with organoids derived from over 100 patients presenting for embryo transfer as described above and compared the levels of hCG production from the culture media with the patient’s clinical history as well as the outcome in the next embryo transfer. When we plotted hCG production levels against the number of previous implantation failures, we found that hCG levels decreased with increasing failure number, with patients who had three or more failures showing uniformly low hCG production (Figure 4A). The difference in average hCG levels between patients with 0-2 previous failures and those with 3 or more was highly significant (p=0.012, Student’s two-sided t-test).

**Figure 4.**
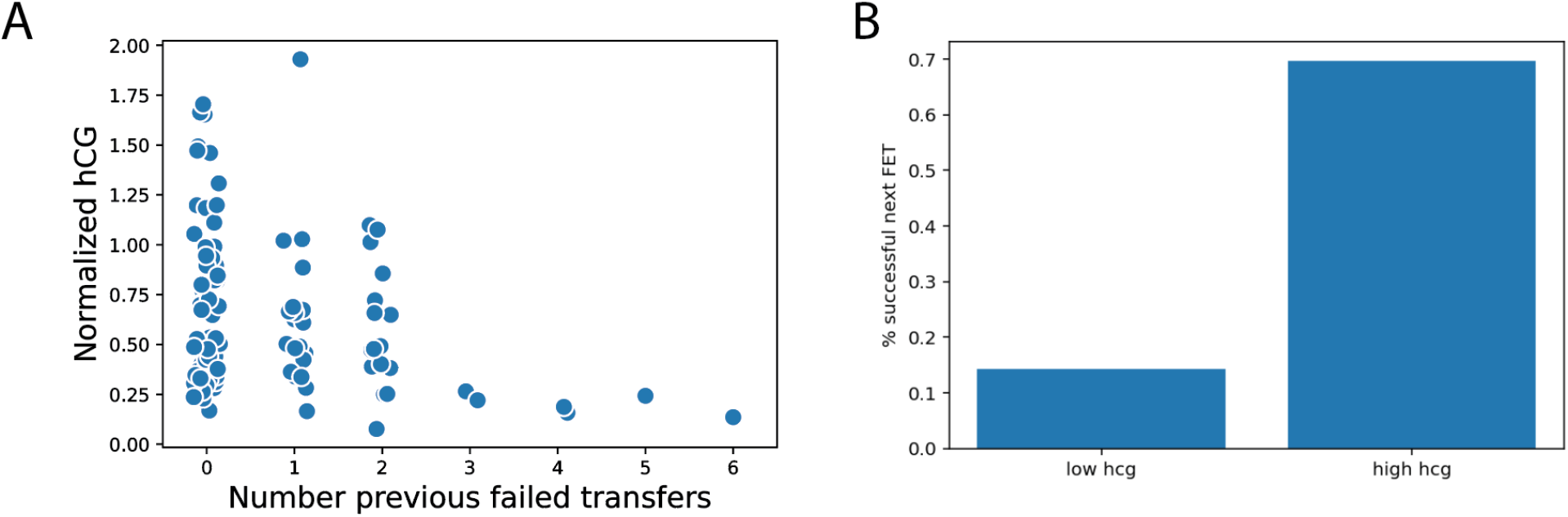
hCG levels induced by organoids correlate with clinical history and outcomes. (A) Levels of hCG are plotted as a function of the number of previous failures. Each data point represents one patient. (B) Patients were subdivided based on the level of hCG production in the organoid-blastoid assay and the outcomes on their next transfer were assessed.

Examining the next embryo transfer, we binarized the data on hCG production and compared the likelihood of success in the low and high bins. We found a large difference in success rates between the two populations with an approximately 10% chance of success in the low hCG population and a 65% chance of success in the hCG high population, comparable to the rates of success in the general population undergoing embryo transfer with euploid embryos (p = 0.006, Fisher’s exact test).

### Including imaging information improves the predictive power of the blastoid-organoid assay

Although the above assay showed a strong correlation with patient outcomes, there were nonetheless some patients with low hCG production who were successful in the next embryo transfer, and we aimed to determine whether imaging the blastoid-organoid interface could provide additional information.. Blastoids created for this study used hPSCs labeled with membrane RFP (RFP::CAAX) to allow for distinguishing the blastoid from the organoid cells. We also immunostained for MUC1, which labeled the organoid cells but not the blastoid cells, and visualized the cytoskeleton and nuclei with phalloidin and DAPI, respectively. We performed confocal imaging of the blastoid-organoid interface for each patient and noted strong variations in the degree to which the blastoid invaded the micropatterned surface displacing the organoid (Figure 5A).

**Figure 5.**
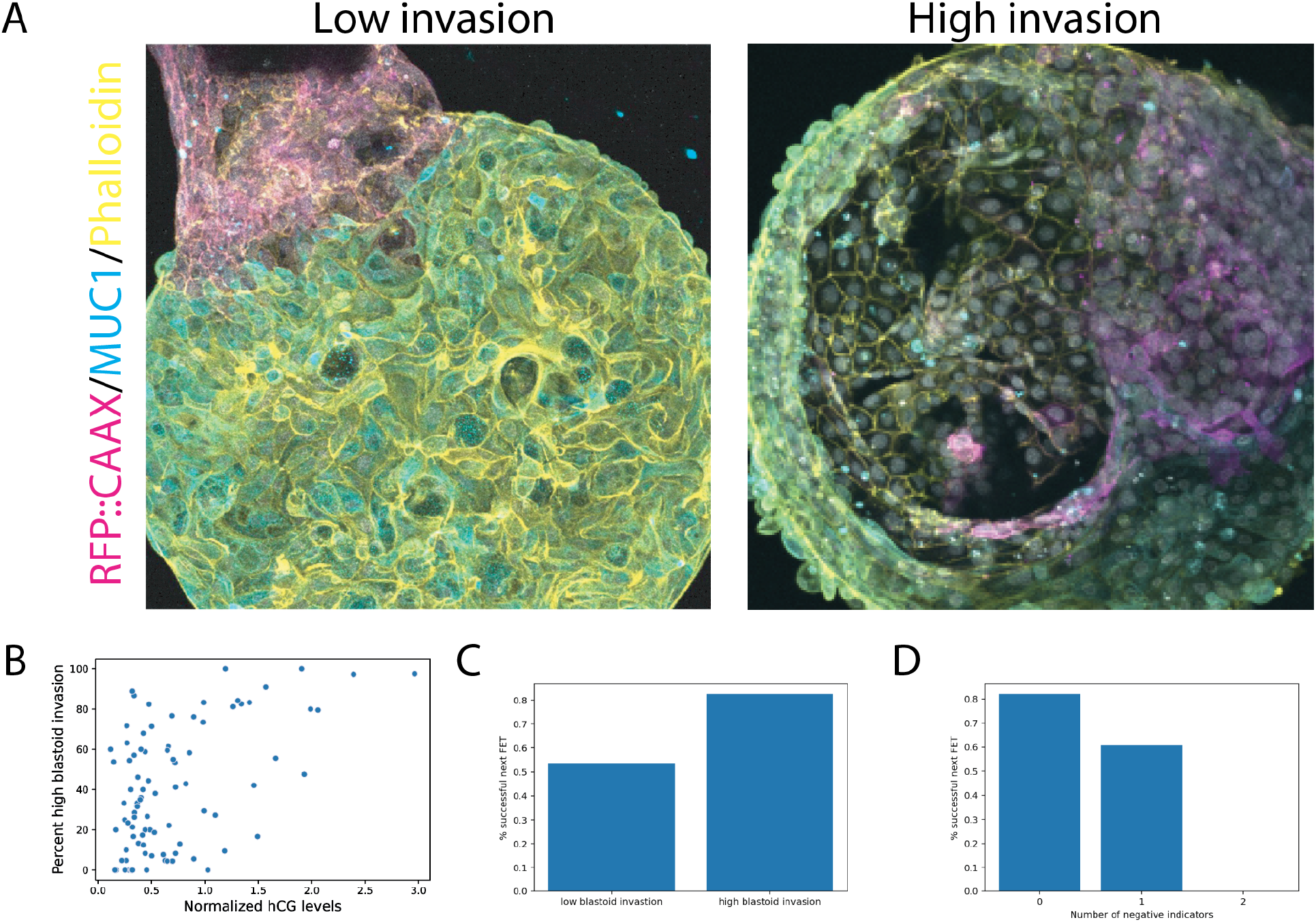
Imaging of the blastoid-organoid interface improves the prediction of clinical outcomes. (A) Example images of high and low blastoid invasion of the micropatterned organoid. (B) Scatterplot comparing the fraction of high invasion organoids with hCG production. Each data point is one patient. (C) Comparison of success rates at the next embryo transfer for patients with a high percentage of high invasion as compared to those with a low percentage. (D) Comparison of success rates on the next embryo transfer as a function of the number of negative indicators from the imaging and hCG measurements.

We hypothesized that organoids which allow blastoids to better attach and expand on the culture surface may be indicative of greater endometrial receptivity in that patient. For each patient, we manually scored each blastoid-organoid interface for the degree of invasion and correlated the fraction of high invasion organoids with clinical outcomes. We first compared the percentage of colonies with high invasion with the levels of hCG production for each patient and found that while they were correlated, there was significant variation (Figure 5B, pearson correlation = 0.51), suggesting that the imaging contained independent information from the hCG measurements. Comparing outcomes with imaging scores, we indeed found a significant difference in success rates between those with a high fraction of high invasion organoids and those with a low fraction (Figure 5C, p = 0.004, Fisher’s exact test).

Finally, we combined the information from imaging and hCG measurements in a simple way. We considered either low hCG or low invasion to be a negative indicator and subdivided the patients based on whether they had 0, 1, or 2 negative indicators. Strikingly, none of the patients with 2 indicators were successful (n = 6, comparison between 0 and 1, p = 0.006, comparison between 0 and 2, p=0.0002, Fisher’s exact test), while those with 0 indicators showed the highest success levels (82% successful, n = 39 patients). Those with 1 indicator were intermediate at 60% success (n = 51, comparison between 1 and 2, p = 0.037 Fisher’s exact test). Thus, incorporating both imaging and hCG measurements showed improved prediction of patient outcomes as compared to measuring either factor alone.

## Discussion

Here we introduced an optimized functional assay involving both imaging and measurement of hCG for simulating the process of implantation in vitro, which we named “Simbryo FX”. We showed that results from this assay correlate with clinical history and with outcomes of a future embryo transfer. We collected biopsy samples from over 100 patients planning for transfer of a euploid embryo and performed transcriptomic measurements of the biopsies and biopsy-derived organoids treated with a cycle of hormones. We then used the organoids in an assay in which blastoids and organoids were co-cultured, hCG levels were measured from the culture media, and imaging was performed to determine the extent to which blastoids invaded the organoids. While transcriptomic measurements did not show any clear relationship with outcomes, both high hCG production and high blastoid invasion as determined by imaging correlated with clinical success. Combining both of these data types yielded the most information about future clinical outcomes.

There are currently no existing tests to directly measure endometrial competence, the degree to which the endometrium of a patient is able to support the process of implantation. Endometrial biopsies are used to measure genes associated with the window of implantation^9,10^, to examine markers of endometriosis^23^ or endometritis^7^, or to measure the microbiome^6^. While each of these tests provides potentially useful information to the patient and clinician, none directly measures endometrial function or the ability to interact with an embryo in order to initiate pregnancy. The test developed here differs from these existing tests in being a functional test of the endometrium that simulates the process of implantation in the lab rather than measuring molecular markers that correlate with different conditions.

The blastoid-organoid test shows promise in predicting clinical outcomes, but there are several areas in which future improvement can be made. The imaging provides a rich dataset offering the possibility of measuring many parameters of the endometrial interaction with the blastoid, however, the current assay uses only the degree of blastoid invasion. The imaging and hCG measurements are combined here in a very simple way, and in the future as more data becomes available, machine learning could combine these measurements, as well as transcriptomics, in order to better predict patient outcomes. Finally, the organoid models utilized here contain only epithelial cells to model the lining of the endometrium, and do not include interactions with the underlying stroma, which play important roles after the initial contact between embryo and endometrium. Using both cell types, as in recently developed assembloids^24–28^, could better capture these interactions.

Here we use this assay as a diagnostic tool to predict the likelihood of success in the next embryo transfer. In the future, it can be used to improve clinical practice. Several protocols can be tried in parallel on the organoids, representing different timings and concentration of hormones or different support agents, and the optimal protocol can be selected based on hCG and imaging measurements. This should allow personalized in vitro optimization of protocols in order to reduce the number of cycles required for a successful transfer.

## Methods

### Study population and clinical procedures

Study participants were recruited from four clinics: Texas Fertility Center (Austin, TX), Aspire HFI (Houston, TX), New Hope Fertility (Houston, TX), and Reach Fertility (Charlotte, North Carolina). The study was approved by WCG IRB (study number 1358882) and participants specifically consented to the procedures performed here including the combination of endometrial organoids with blastoids. Deidentified information on patient histories and outcomes was provided to Simbryo by the clinics using standardized case report forms.

Biopsies were obtained from a total of 139 patients. Of these, RNA was obtained from 119 primary samples with the remainder of samples not yielding sufficient RNA for sequencing (18 samples) or subsequently found to derive from a patient with a blood borne pathogen (2 samples). The data from one sample failed quality control and were not included in the analysis. Of the 118 samples included in the RNA sequencing analysis, 102 participants were reported by the clinic to have transferred a euploid embryo and the outcome was given. Organoids were able to be grown from the samples of 111 participants. Of these, sequencing data was obtained from the organoids with or without hormones from 101 samples and the blastoid-organoid assay was run on all 111 samples. Outcomes were reported for 86/101 and 96/111 samples from the RNA sequencing and blastoid-organoid assay, respectively.

All participants had at least one euploid embryo with intent to transfer and were undergoing a biopsy as part of their routine care. Biopsies were collected using a pipelle endometrial suction curette. In cases where the patient was undergoing a hysteroscopy, biopsies were collected using a pipelle prior to installation of fluid for the hysteroscopy. Biopsies were transported on ice in collection medium (RPMI 1640 (Thermo Fisher), 1% penicillin-streptomycin (Thermo Fisher), 1× GlutaMAX, and 10 mM HEPES) to the laboratory for processing.

### Endometrial biopsy sample collection

Upon receipt in the laboratory, endometrial tissue was washed in cold PBS to remove blood and mucus, then dissected into portions. Samples were transferred into 15 mL conical tubes containing cold DMEM/F-12, centrifuged at 500xg for 5 minutes, and resuspended in cold cryopreservation medium (DMEM/F-12 supplemented with 40% FBS, 10% DMSO, and 10 µM RI), and frozen in a controlled-rate freezing container at −80°C overnight before being transferred to long-term cryostorage.

### Thawing of frozen biopsy samples

Frozen biopsy samples were thawed by placing cryovials in a 37°C water bath for approximately 1.5 minutes until just thawed. The contents were immediately transferred to a 15 mL conical tube containing 10 mL of cold DMEM/F-12 and centrifuged at 500xg for 3 minutes. The supernatant was removed, and the tissue pellet was used directly for organoid generation or RNA extraction.

### Human endometrial organoid (hEMO) generation

Endometrial organoids were generated following the standard protocols developed by Turco et al. 2017^12^ and Boretto et al. 2017^13^ with slight modifications. Endometrial tissue in 1 mL DMEM/F-12 was minced into very small pieces (0.5 mm3) in a 35 mm dish with a dissecting blade. 1 mL collagenase IV (Thermo Fisher, 2 mg/mL) was then added and the dish was incubated at 37°C for 1.5 hours. The tissue and medium were pipetted up and down 50 times with a plastic pasteur pipette every 30 minutes to aid the digestion. The digestion was stopped by adding 2 mL DMEM/F-12 when there were enough free glandular elements by visual inspection. The entire digested sample was collected and transferred into a 15 mL conical tube, and allowed to stand for 2 minutes.

For organoid generation, the tube was centrifuged at 500xg for 3 minutes, and the supernatant was discarded. The pellet was washed once with 10 mL of DMEM/F-12. Matrigel (Corning) was added in the amount of 10x the estimated pellet volume, and the tube was placed on ice. The glandular elements were mixed well with the Matrigel prior to dropping 20-25 μL of Matrigel/cell suspension in the center of wells of 48-well tissue culture plates. The plate was immediately placed upside down in the 37°C incubator. After the Matrigel set (~10 minutes), each drop was overlayed with 250 μL of hEMO Medium (DMEM/F-12, 1% penicillin-streptomycin, 1× GlutaMAX, 1× N2, 1× B27, 1× insulin-transferrin-selenium (Thermo Fisher), 2 ng/mL FGF-2, 50 ng/mL EGF (Thermo Fisher), 10 ng/mL HGF (Thermo Fisher), 10 ng/mL FGF-10 (Thermo Fisher), 500 nM A83-01, 10 µM SB202190 (MCE), 1.25 mM N-acetylcysteine (MCE), 2.5 mM nicotinamide (Thermo Fisher), 100 ng/mL Noggin (Thermo Fisher), and 80 ng/mL RSPO1 (BioTechne)). Medium was changed every other day and organoids were grown for 7-10 days before the first passage.

### Endometrial organoid culture

Throughout the entire passaging procedure, the tubes were kept on ice, and all centrifugation steps were carried out at 4°C. To passage the organoids (following the protocol from Turco et al. 2017, with slight modifications), culture medium was first removed from each well and 250 μL of cold DMEM/F-12 was added into the wells. The matrigel drop was detached into the medium by scraping backwards and forwards across the surface of each well by using a 1 mL pipette tip. The organoid suspension was also pipetted up and down to help break the Matrigel and the organoids. The contents of three to five wells were pooled into one low-adhesion 2 mL microcentrifuge tube. The tube was centrifuged at 500xg for 3 minutes to pellet the organoids. The supernatant was removed, and 500 μL of cold DMEM/F-12 was added to each tube.

The organoids and Matrigel were then disrupted by pipetting up and down 200 times through a 200 μL pipette tip. Following this, 1 mL of DMEM/F-12 was added, and centrifugation was performed. The supernatant was removed, and 500 μL of DMEM/F-12 was again added, followed by further manual pipetting up and down 100 times. Another 1 mL of DMEM/F-12 was added, and centrifugation was performed again at 500xg for 4 minutes. After removing the supernatant, the pellet was re-suspended gently in Matrigel. Each original well should be split into two or three wells. Droplets of 20-25 μL of Matrigel/cell suspension were pipetted onto the center of the wells in a 48-well culture plate. The culture plate was placed upside down in the 37°C incubator for 10 minutes to allow the Matrigel to set. Finally, 250 μL of hEMO medium was added to each well. The medium was changed every other day, and passaging was carried out every 7-10 days.

### Implantation assay

Implantation assay was performed as described in Khoei et al. 2023^18^ with some modifications. Endometrial organoids at passage five were used for the implantation assay. On Day 0, organoids were passaged as previously described and seeded into 32 wells of a 48-well plate. On Day 2, the medium was changed. On Day 4, 14 and 18 random wells were assigned for treatment with DMSO and hormones, respectively. Medium in hormone-assigned wells were changed into hEMO medium supplemented with 10 nM Estradiol/E2 (MCE) on Day 4-5, followed by hEMO medium supplemented with EPCX (10 nM E2, 1 µM progesterone (MCE), 250 µM 8-bromo-cAMP (MCE), and 2 µM XAV-939) on Day 6-7. Medium in control-assigned wells were replaced with hEMO medium supplemented with DMSO.

On Day 8, organoids were passaged to single cells. Passaging procedure was performed as previously described, but instead of manual pipetting for 200 times, organoids were incubated with TrypLE Express for 7 minutes. Dissociation was stopped by adding cold DMEM/F-12, followed by centrifugation. After the supernatant was removed, the cell pellet was mixed up and down 100 times in 200 µL cold DMEM/F-12. The cell suspension was then filtered through a 40 µm filter to obtain single cells. 35,000 cells were seeded in a well of a 96-well micropattern (CYTOO) plate coated with 3% Matrigel the day before. Prior to seeding, these wells were washed five times with cold DMEM/F-12 to remove extra Matrigel. Three wells were used per condition. The plate was incubated at 37°C for 3.5 hours, and the wells were washed four times with warm DMEM/F-12 to remove cells that did not attach to the patterns. After the final wash, hEMO medium supplemented with DMSO or EPCX was added into the wells. On Day 9-10, medium was replaced with hEMO medium supplemented with DMSO or EPCX.

On Day 11, cells were washed twice with warm DMEM/F-12, and CMRL-1 medium (CMRL (Thermo Fisher) medium supplemented with 10% embryonic stem-cell FBS (Thermo Fisher), 2 mM L-glutamine, 1× N2, 1× B27, 1 mM sodium pyruvate, 10 nM E2, 1 µM progesterone, and 10 µM RI) was added. Cells were incubated in CMRL-1 medium for at least 2 hours.

Meanwhile, 900 µL of medium was removed from each well of the Aggrewell plate. To remove blastoids from the well, 400 µL of warm N2B27 was gently added into the well and blastoids were pipetted up and down very gently using a 200 µL wide bore tip. The blastoids were transferred into a 35 mm dish containing 1.5 mL of N2B27. Plates and dishes were kept warm at 37°C by using a heating pad. Blastoids with good morphology (i.e., correct size, presence of inner cell mass) were selected by using a stereomicroscope. Ten blastoids were transferred into each well of a 96-well microplate with cell-repellent surface (Greiner) containing 100 µL of N2B27. Once blastoids selection was finished, 70 µL of the medium was removed from each well and replaced with 70 µL of CMRL-1 medium. Everything was transferred into the wells containing the endometrial cells and the plate was incubated at 37°C in hypoxic conditions. After 2 days, hCG measurement and immunostaining were performed.

### Human Chorionic Gonadotropin (hCG) measurement

Measurement of hCG was performed by using a b-hCG ELISA kit (DRG International) following the manufacturer’s protocol. In short, 25 µL of cell culture media from control and hormone-treated wells were dispensed onto the wells of the ELISA plate, followed by 100 µL of the enzyme conjugate solution, and incubated for one hour. The plate was then washed five times with distilled water, and 100 µL of substrate solution was added. After 15 minutes, 50 µL of the stop solution was added. The absorbance at 450 nm and 630 nm was read with a standard plate reader.

### Immunostaining

Immunostaining was performed at room temperature according to the following. The samples were washed once in PBS and fixed with 4% paraformaldehyde for 20 minutes. The samples were then washed twice in PBS, and blocked with 3% donkey serum and 0.1% Triton-X in PBS for 30 minutes. After removing the blocking solution, the primary antibody diluted in the blocking solution was added and incubated for two hours. The samples were then washed four times for a total of 30 minutes with PBST (PBS + 0.1% Tween-20). Secondary antibodies (1:500), DAPI (1:500), and/or phalloidin (1:100) diluted in the blocking solution were added and incubated for 30 minutes. The samples were then washed once with PBST for 30 minutes, followed by two more washes with PBST for 5 minutes each. The samples were stored in PBS at 4°C, sealed with parafilm. Imaging was performed with a laser scanning confocal microscope with 20x objective magnification.

### RNA extraction and sequencing

RNA extraction from endometrial biopsies followed the same protocol as described for hEMO generation, with modifications after the digestion step. Following digestion, all material was collected and transferred into a 15 mL conical tube, centrifuged at 500xg for 3 minutes, and the supernatant was removed. The cell pellet was resuspended in 700 µL of lysis buffer and incubated for 45 minutes to ensure complete lysis. The lysate was then transferred to a homogenizer column (Thermo Fisher) and centrifuged at 12,000xg for 2 minutes. The resulting flow-through was used for RNA purification using the PureLinkTM RNA Mini Kit (Thermo Fisher), following the manufacturer’s instructions. Purified RNA was either stored at −80°C or submitted to Novogene for sequencing.

Endometrial organoids were prepared following the same protocol used for the implantation assay, except that cultures were established in regular (non-micropatterned) plates. Instead of proceeding with the implantation assay, RNA was extracted using the PureLinkTM RNA Micro Kit (Thermo Fisher) according to the manufacturer’s instructions. Purified RNA was either stored at −80°C or submitted to Novogene for sequencing.

### RNA-seq data analysis

Transcript-level abundance was quantified using Salmon (v1.10.3) in quasi-mapping mode against the human transcriptome reference (GRCh38 cDNA). Transcript quantifications were imported and annotated using tximeta (v1.22.1) in R (v4.4.1), and summarized to gene-level counts for downstream analysis. All downstream analysis, including normalization and differential gene expression, were performed using standard workflows with custom code written in Python.

### Quantitative RT-PCR

Total RNA was extracted using the PureLinkTM RNA Micro Kit (Thermo Fisher) according to the manufacturer’s instructions. Complementary DNA (cDNA) was synthesized using the High-Capacity cDNA Reverse Transcription Kit (Thermo Fisher). Quantitative PCR (qPCR) was performed using the Luna® Universal qPCR master mix (New England Biolabs) on a Quant Studio 5 Real-Time PCR system (Thermo Fisher). GAPDH expression was used as an internal control.

## Data Availability

All data produced in the present study are available upon reasonable request to the authors

## Acknowledgements

We thank Dr Elena Camacho Aguilar for early experiments with endometrial organoids and Dr David Huang (UCSF) for a critical reading of the manuscript.

## References

1. Gaskins, A. J., Zhang, Y., Chang, J. & Kissin, D. M. Predicted probabilities of live birth following assisted reproductive technology using United States national surveillance data from 2016 to 2018. American Journal of Obstetrics and Gynecology 228, 557.e1–557.e10 (2023).

2. Sun, Y., Zhang, Y., Ma, X., Jia, W. & Su, Y. Determining Diagnostic Criteria of Unexplained Recurrent Implantation Failure: A Retrospective Study of Two vs Three or More Implantation Failure. Front. Endocrinol. 12, 619437 (2021).

3. Pirtea, P. et al. Recurrent implantation failure: reality or a statistical mirage? Fertility and Sterility 120, 45–59 (2023).

4. Pirtea, P. et al. Rate of true recurrent implantation failure is low: results of three successive frozen euploid single embryo transfers. Fertility and Sterility 115, 45–53 (2021).

5. Smith, A. D. A. C., Tilling, K., Nelson, S. M. & Lawlor, D. A. Live-Birth Rate Associated With Repeat In Vitro Fertilization Treatment Cycles. JAMA 314, 2654 (2015).

6. Moreno, I. et al. Evidence that the endometrial microbiota has an effect on implantation success or failure. American Journal of Obstetrics and Gynecology 215, 684–703 (2016).

7. Moreno, I. et al. The diagnosis of chronic endometritis in infertile asymptomatic women: a comparative study of histology, microbial cultures, hysteroscopy, and molecular microbiology. American Journal of Obstetrics and Gynecology 218, 602.e1–602.e16 (2018).

8. Toson, B., Simon, C. & Moreno, I. The Endometrial Microbiome and Its Impact on Human Conception. IJMS 23, 485 (2022).

9. Díaz-Gimeno, P. et al. A genomic diagnostic tool for human endometrial receptivity based on the transcriptomic signature. Fertility and Sterility 95, 50–60.e15 (2011).

10. Ohara, Y. et al. Clinical relevance of a newly developed endometrial receptivity test for patients with recurrent implantation failure in Japan. Reprod Medicine & Biology 21, e12444 (2022).

11. Doyle, N. et al. Effect of Timing by Endometrial Receptivity Testing vs Standard Timing of Frozen Embryo Transfer on Live Birth in Patients Undergoing In Vitro Fertilization: A Randomized Clinical Trial. JAMA 328, 2117 (2022).

12. Turco, M. Y. et al. Long-term, hormone-responsive organoid cultures of human endometrium in a chemically defined medium. Nat Cell Biol 19, 568–577 (2017).

13. Boretto, M. et al. Development of organoids from mouse and human endometrium showing endometrial epithelium physiology and long-term expandability. Development dev.148478 (2017) doi:10.1242/dev.148478.

14. Boretto, M. et al. Patient-derived organoids from endometrial disease capture clinical heterogeneity and are amenable to drug screening. Nat Cell Biol 21, 1041–1051 (2019).

15. Kagawa, H. et al. Human blastoids model blastocyst development and implantation. Nature 601, 600–605 (2022).

16. Karvas, R. M. et al. 3D-cultured blastoids model human embryogenesis from pre-implantation to early gastrulation stages. Cell Stem Cell 30, 1148–1165.e7 (2023).

17. Liu, X. et al. Modelling human blastocysts by reprogramming fibroblasts into iBlastoids. Nature 591, 627–632 (2021).

18. Khoei, H. H. et al. Generating human blastoids modeling blastocyst-stage embryos and implantation. Nat Protoc 18, 1584–1620 (2023).

19. Altmäe, S. et al. Meta-signature of human endometrial receptivity: a meta-analysis and validation study of transcriptomic biomarkers. Sci Rep 7, 10077 (2017).

20. Takashima, Y. et al. Resetting transcription factor control circuitry toward ground-state pluripotency in human. Cell 158, 1254–1269 (2014).

21. Guo, G. et al. Human naive epiblast cells possess unrestricted lineage potential. Cell Stem Cell 28, 1040–1056.e6 (2021).

22. Linneberg-Agerholm, M. et al. Naïve human pluripotent stem cells respond to Wnt, Nodal and LIF signalling to produce expandable naïve extra-embryonic endoderm. Development 146, dev180620 (2019).

23. Evans-Hoeker, E. et al. Endometrial BCL6 Overexpression in Eutopic Endometrium of Women With Endometriosis. Reprod. Sci. 23, 1234–1241 (2016).

24. Rawlings, T. M. et al. Modelling the impact of decidual senescence on embryo implantation in human endometrial assembloids. eLife 10, e69603 (2021).

25. Shibata, S. et al. Modeling embryo-endometrial interface recapitulating human embryo implantation. Sci. Adv. 10, eadi4819 (2024).

26. Song, J. et al. 3D post-implantation co-culture of human embryo and endometrium. Cell Stem Cell 33, 58–72.e7 (2026).

27. Li, Q. et al. A 3D in vitro model for studying human implantation and implantation failure. Cell 189, 70–86.e20 (2026).

28. Molè, M. A. et al. Modeling human embryo implantation in vitro. Cell 189, 87–105.e28 (2026).

