## Supplementary Figures 1-7 for "A novel functional assay based on patient-derived endometrial organoids and blastoids predicts the success of embryo transfer"

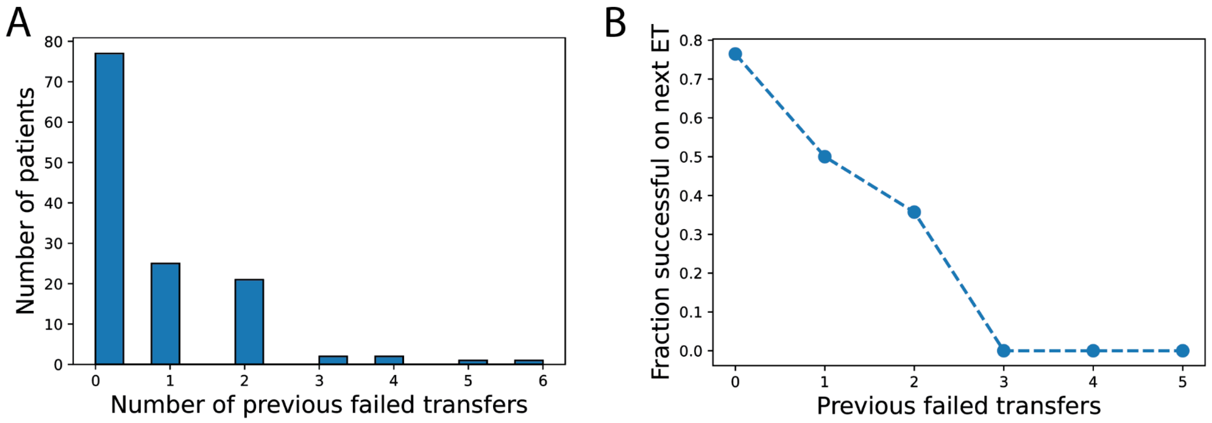


**Figure S1. Previous and next transfer success in the study population.** (A) Distribution of number of previous failed transfers for patients participating in the study. (B) Fraction of patients successful in the next transfer of a euploid embryo as a function of the number of previous failures.

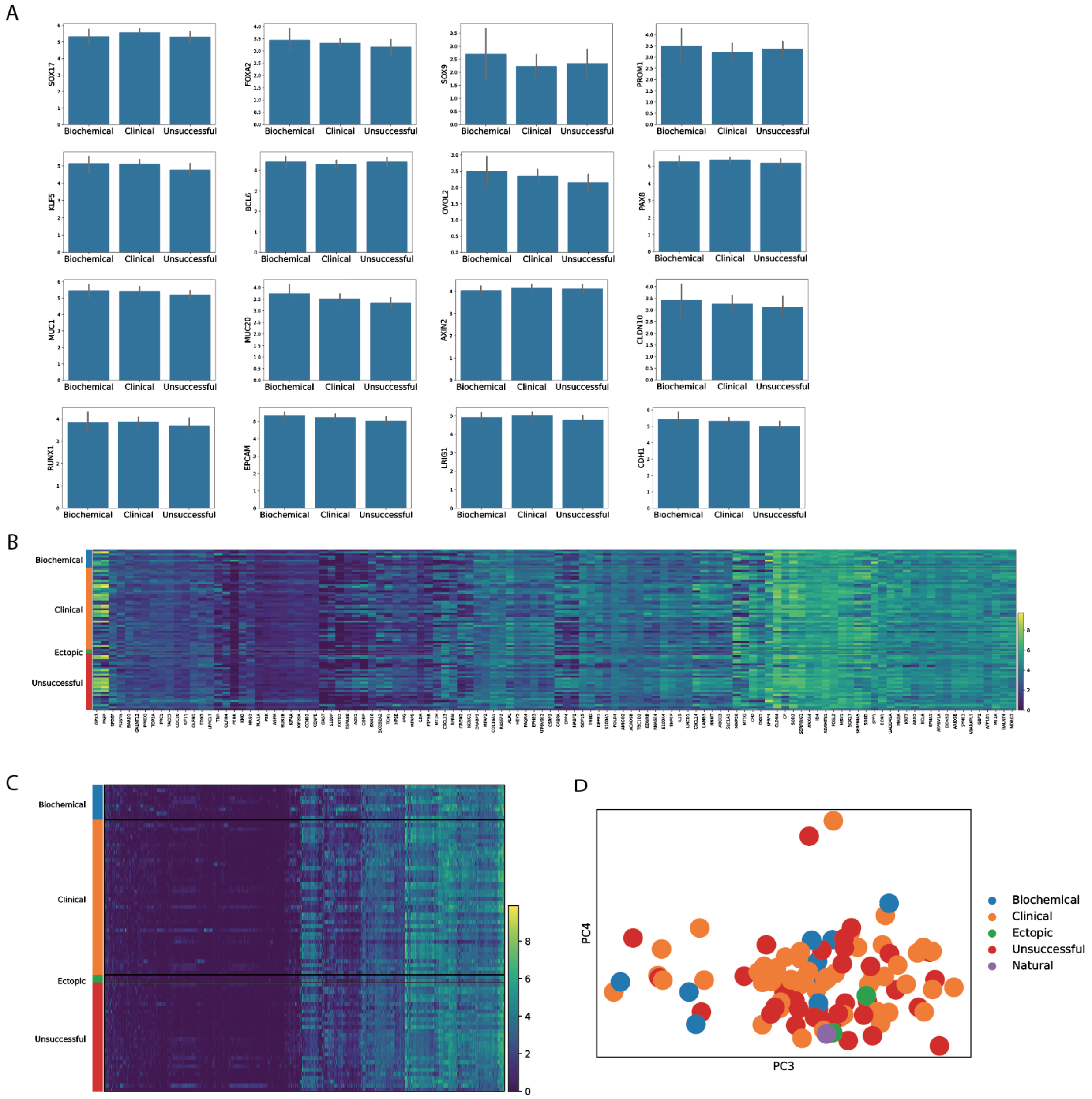


**Figure S2. Correlations between gene expression in the primary biopsy samples and clinical outcomes.** (A) Comparison of average expression of known endometrial marker genes in patients with different clinical outcomes (biochemical pregnancy, clinical pregnancy, unsuccessful transfer) (B) Expression of genes used in the ERA assay in primary samples from patients. Patients are grouped based on their clinical outcomes as indicated on the left. (C) As in B except data is shown for the 2000 most highly variable genes in the dataset. (D) Plot of PC3 vs PC4 color coded by clinical outcomes.

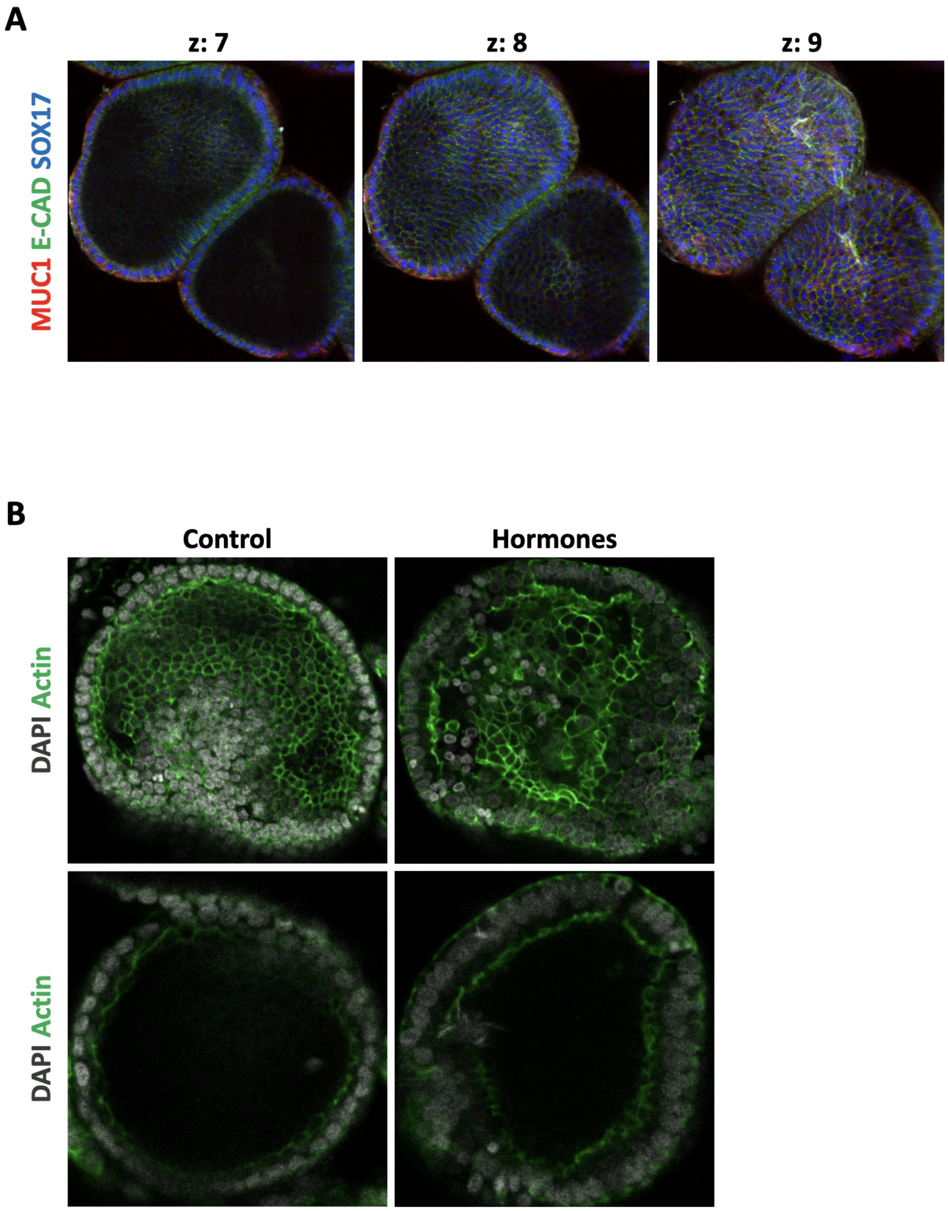


**Figure S3. Imaging of endometrial organoids.** (A) Images of several z-slices of an organoid stained with the indicated markers. (B) Comparison of the morphology of organoids with and without treatment with a cycle of hormones. Actin cytoskeleton and nuclei are stained with phalloidin and DAPI, respectively.

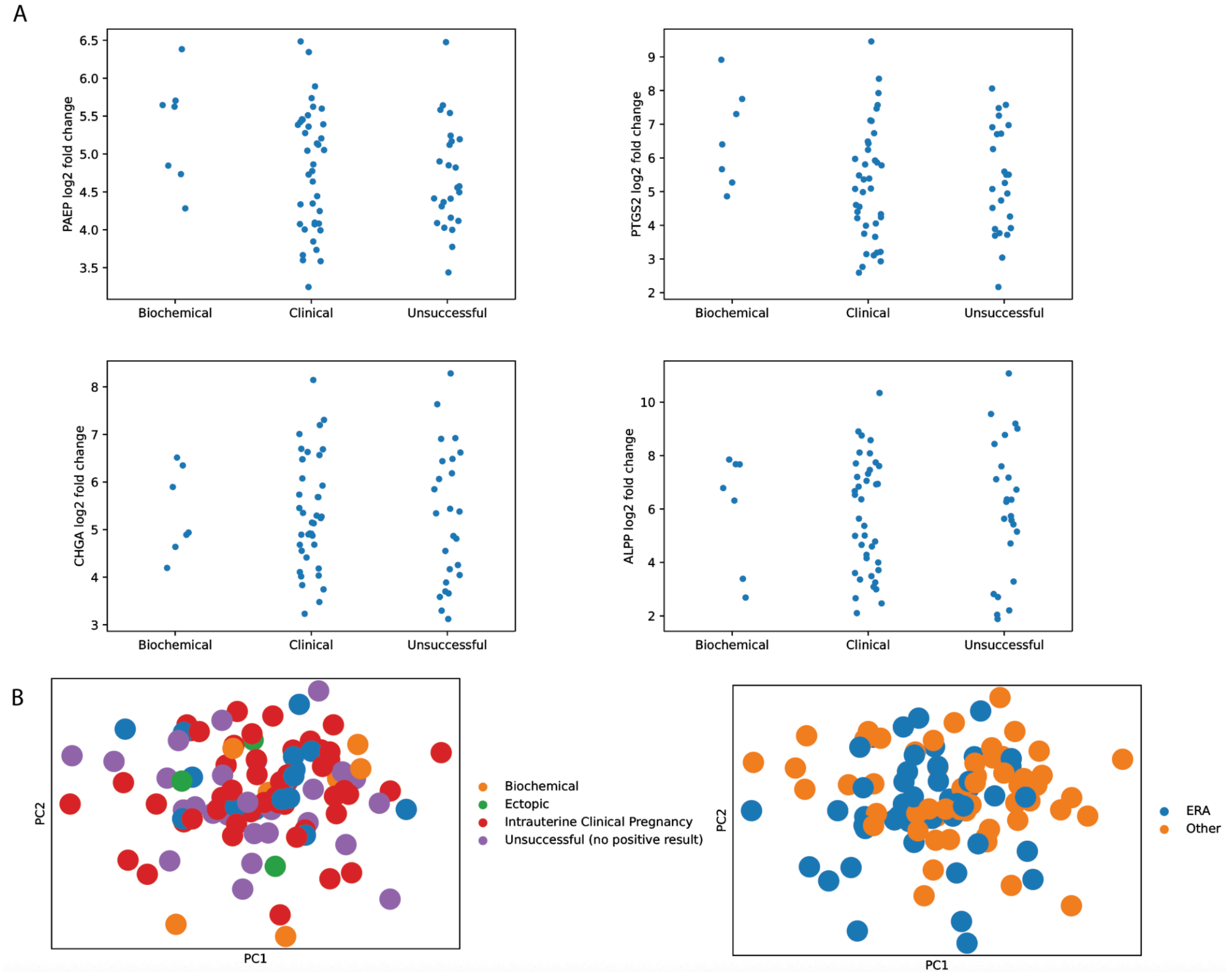


**Figure S4. Correlation of hormone response in organoids with clinical outcomes.** (A) log2 fold change of indicated progesterone responsive genes separated by clinical outcome. Each data point is one patient. (B) PCA plot generated from the matrix of fold changes induced by hormones. The plot is color coded by clinical outcomes (left) or by whether patients underwent the biopsy for a window of implantation test (right)..

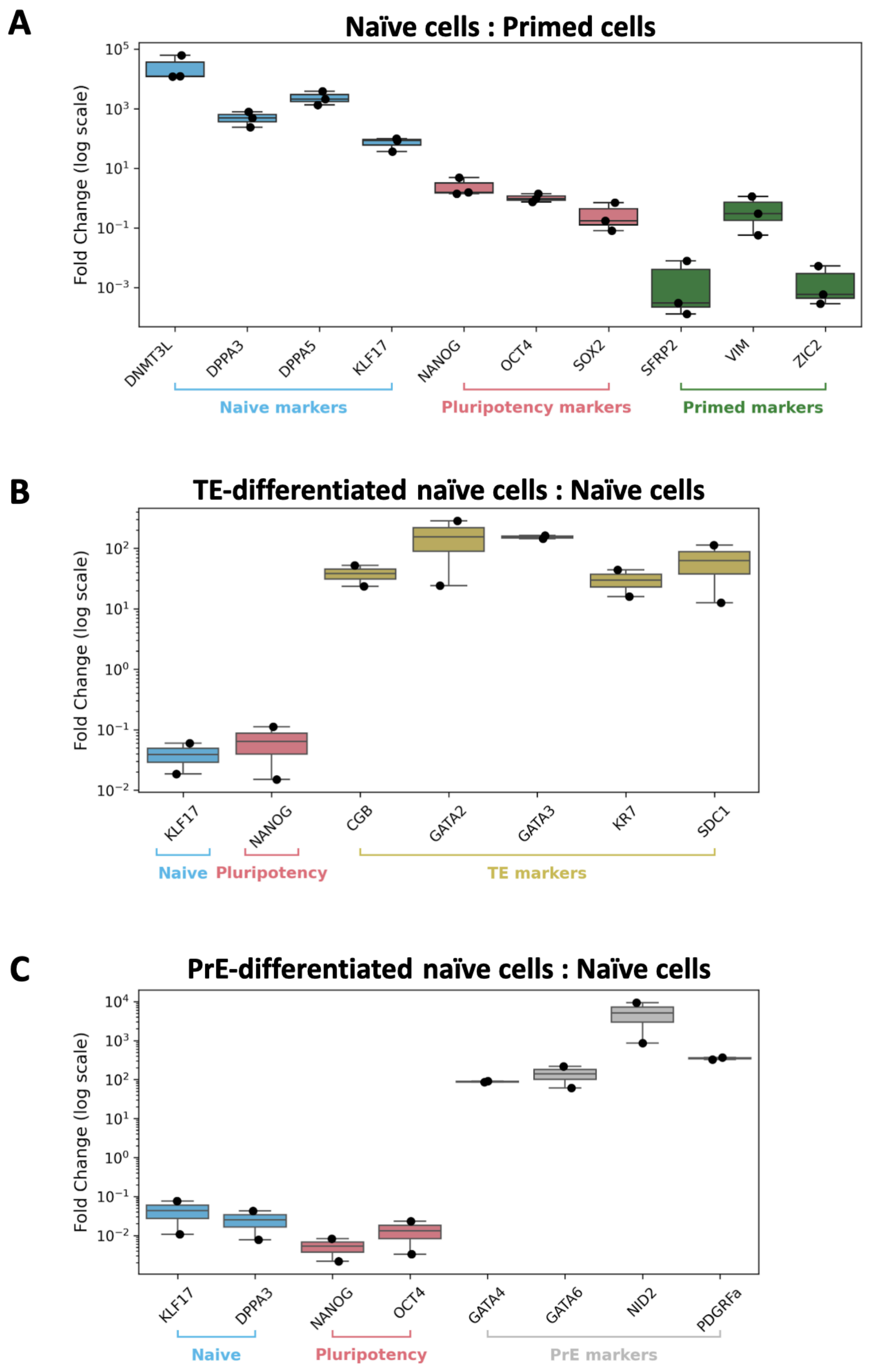


**Figure S5. Gene expression measurements in pluripotent and differentiated cells.** (A-C) Fold changes for the indicated markers as determined by qPCR between (A) naive and primed PSCs (B) differentiated TE and naive PSCs (C) differentiated PrE and naive PSCs.

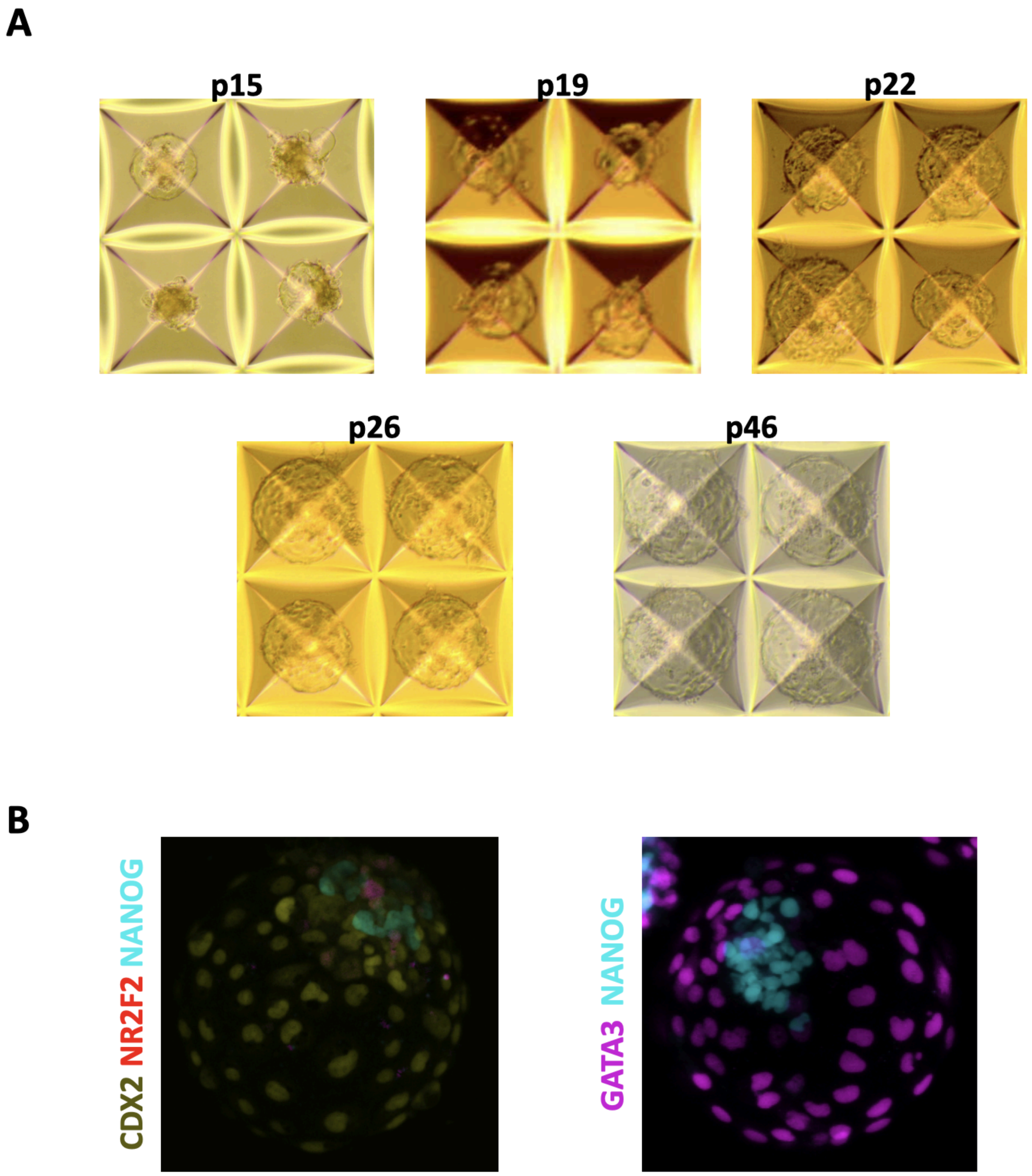


**Figure S6. Blastoids generated from naive cells.** (A) Example images of blastoids generated after the indicated number of passages in the naive state. (B) Images of blastoids stained for the indicated markers.

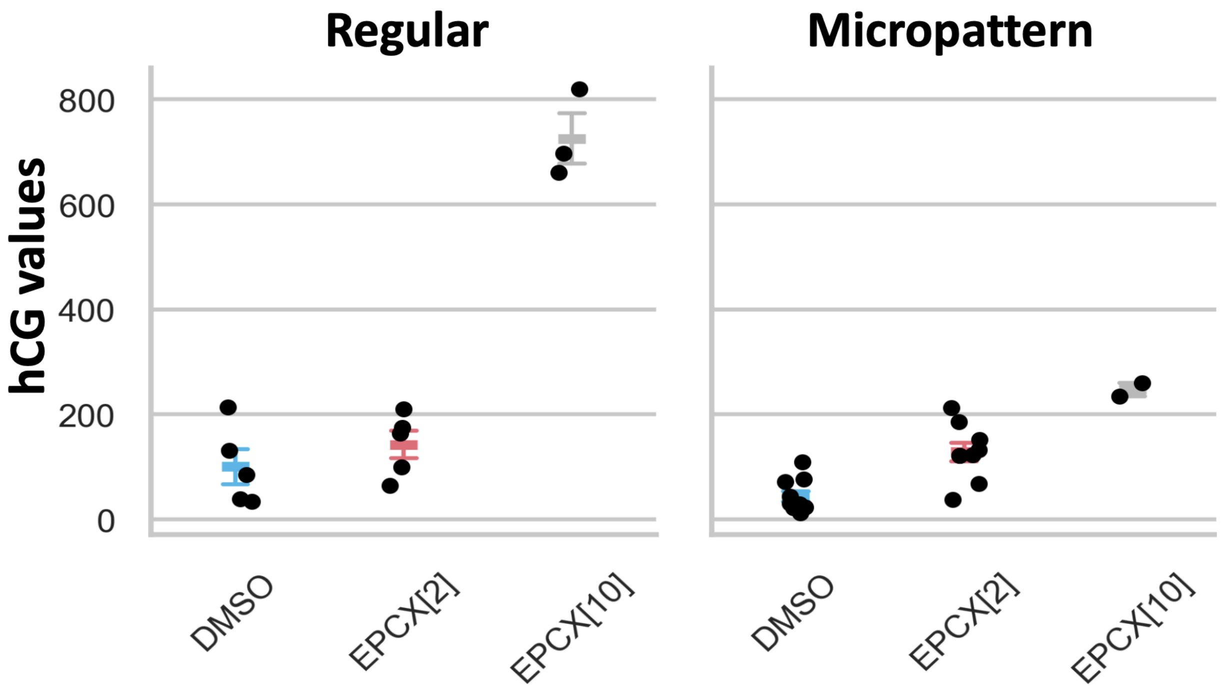


**Figure S7. Hormone production by blastoids without endometrial organoids.** Blastoids were grown either with or without hormones (EPCX). The number in parentheses indicates the concentration of the XAV939 Wnt inhibitor in uM. 2 uM was used throughout this study.
